# Development and evaluation of Z-score based aortic diameter thresholds for early detection of thoracic aortic dissection and aneurysm: Analysis in the UK Biobank

**DOI:** 10.1101/2025.10.03.25337259

**Authors:** Nikhil Paliwal, Albert Henry, Chris Finan, Rhodri H. Davies, R. Thomas Lumbers, Janice Tsui, Alun D. Hughes, Bryan Williams, Aroon D. Hingorani

## Abstract

**Background & Purpose:** Clinical guidelines recommend using an absolute ascending aortic diameter (AAD) cutoff of 4.5 cm for monitoring and 5.5 cm for surgery in people at risk of thoracic aortic dissection. However, AAD varies with age, sex, height and body surface area (BSA). Using imaging data from the UK Biobank, we investigated different AAD cutoffs in the prediction of aortic dissection or aneurysm based on normal population variation in AAD, with and without adjustment for age, sex, height and BSA.

**Methods:** Using cardiac magnetic resonance (CMR) images of UK Biobank participants, we applied a pre-trained neural network to segment and measure AAD. We evaluated four approaches to predict a subsequent clinical diagnosis of thoracic aortic dissection or presence of a thoracic aneurysm. We evaluated (i) the performance of the guideline endorsed AAD cutoff of 4.5 cm, and (ii) cutoffs based on the standardized deviation from the population mean aortic diameter (Z-score): a) without accounting for age, sex or anthropometric measures (Z_AAD_); b) accounting for age, height, and sex (Z_H_) and c) accounting for age, body surface area (BSA) and sex (Z_BSA_). We assessed performance using the detection rate (DR) and false positive rate (FPR).

**Results:** We measured AAD from CMR images of 77,527 participants from UK Biobank, among whom there were 72 subsequent diagnoses of thoracic dissection or aneurysm (incidence rate ∼0.1%, approximately 1 in 1100). The AAD was 4.2 cm (SD 0.6) in affected individuals and 3.3 cm (SD 0.4) in unaffected individuals (p<0.001). The guideline endorsed 4.5 cm absolute AAD cutoff had a DR of 25% at a 0.3% FPR. A cutoff of Z_AAD_>2 detected 57% of affected individuals at a 3% FPR, while Z_H_>2 and Z_BSA_>2 had DRs of 53% and 47% respectively, with FPRs of 4% and 2%, respectively. The area under the ROC curve (AUC) was 0.90 for Z_AAD,_ 0.84 for Z_H_ and 0.83 for Z_BSA_.

**Conclusion:** The current guideline-recommended absolute AAD cutoff of 4.5 cm has a low FPR but a low DR, missing nearly three-quarters of individuals who later developed thoracic aneurysm or dissection. Z-score-based approaches, particularly the Z_AAD_, improve the DR at the expense of a higher FPR. These findings support the consideration of population-derived Z-scores in clinical monitoring and risk stratification for thoracic aortas at risk of dissection.

## INTRODUCTION

Thoracic aortic dissection has an annual incidence of 3–4 per 100,000 people, and an in-hospital mortality rate approaching 30%.(1–3) Increasing aortic diameter is associated with a higher risk of aortic dissection.(4) Many individuals with thoracic aortic enlargement remain undiagnosed, and aortic dilation or aneurysm is often detected incidentally during cardiac imaging undertaken for other reasons, such as evaluation of left ventricular wall thickness in people with hypertension. Recent clinical guidelines from the European Society of Cardiology (ESC) and American Heart Association (AHA) recommends monitoring aortic diameter with absolute cut-offs, typically 4.5 cm for surveillance and 5.5 cm for consideration of prophylactic surgery in non-genetic cases.(5–7) However, the International Registry of Acute Aortic Dissections (IRAD) has established that more than half of dissected thoracic aortas had diameters less than 5.5 cm.(2)

Aortic diameter varies in the population and is associated with age, sex, height and body surface area.(8) Therefore, an approach based on the distribution of AAD values in the population (a Z-score based approach) has been proposed as an alternative to a single absolute AAD value for monitoring or surgical intervention, including incorporating adjustments for age, sex and body habitus. For example, Z-scores adjusted for height and body surface area (BSA) (see equations 1 and 2 below) have been proposed and evaluated with a cut-off of Z>2 SD being used for the designation of high risk of thoracic aortic dissection.(9) Z-scores are also recommended in populations with smaller stature, such as Turner syndrome, as endorsed in ESC guidelines.(5, 6, 10) A study by van Kimmenade et al. compared Z-scores derived from different adjustment methods (age, body surface area, and height) in patients with Marfan syndrome, showing that height-based adjustment provided the highest accuracy.(9) However, the performance of cutoffs based on the standardized deviation from the population mean aortic diameter (Z-score) (with and without adjustment for other variables) has not previously been evaluated.

We used cardiac magnetic resonance (CMR) imaging of the aorta in participants in UK Biobank (UKB) to compare the performance of a single absolute AAD cut-off of 4.5cm with a Z-score based approach in the prediction of a later diagnosis of thoracic aortic aneurysm or dissection. In addition to previously reported Z-scores adjusted for BSA or height, we used an unadjusted Z-score based simply on the distribution of AAD values in the UKB.

## METHODS

### Study Population

UKB is a richly phenotyped, prospective population cohort with over half a million participants, and a subset receiving CMR imaging.(11, 12) Data was accessed under application number 12113 from the University College London. UK Biobank received approval from the National Information Governance Board for Health and Social Care and the National Health Service North West Centre for Research Ethics Committee (Ref: 11/NW/0382). All participants supplied written consent for the use of their data. MRI was conducted during the third visit of each participant using a Siemens 1.5 Tesla MAGNETOM Aera scanner (Siemens Healthineers, Erlangen, Germany). Details of the imaging protocol can be found in Petersen et al.(11) We downloaded and processed the aortic cine images (UKB field 20210) from the UKB bulk downloads for 77,527 participants.

For participants with an aortic image, we also extracted common clinical and demographic covariates like age, sex, body mass index (BMI), height, weight, systolic blood pressure (SBP) and diastolic blood pressure (DBP). Using the height and weight, we calculated body surface area (BSA) for each participant using the Dubois formula.(13)

### Image Segmentation & AAD Calculation

Aortic CMR images were automatically segmented using a validated pre-trained neural network.(14, 15) After aortic segmentation, maximum aortic area was identified and was used to evaluate the aortic diameter assuming a circular aortic cross-section. Figure 1 shows a flowchart with an overview of the calculation of AAD.

**Figure 1.**
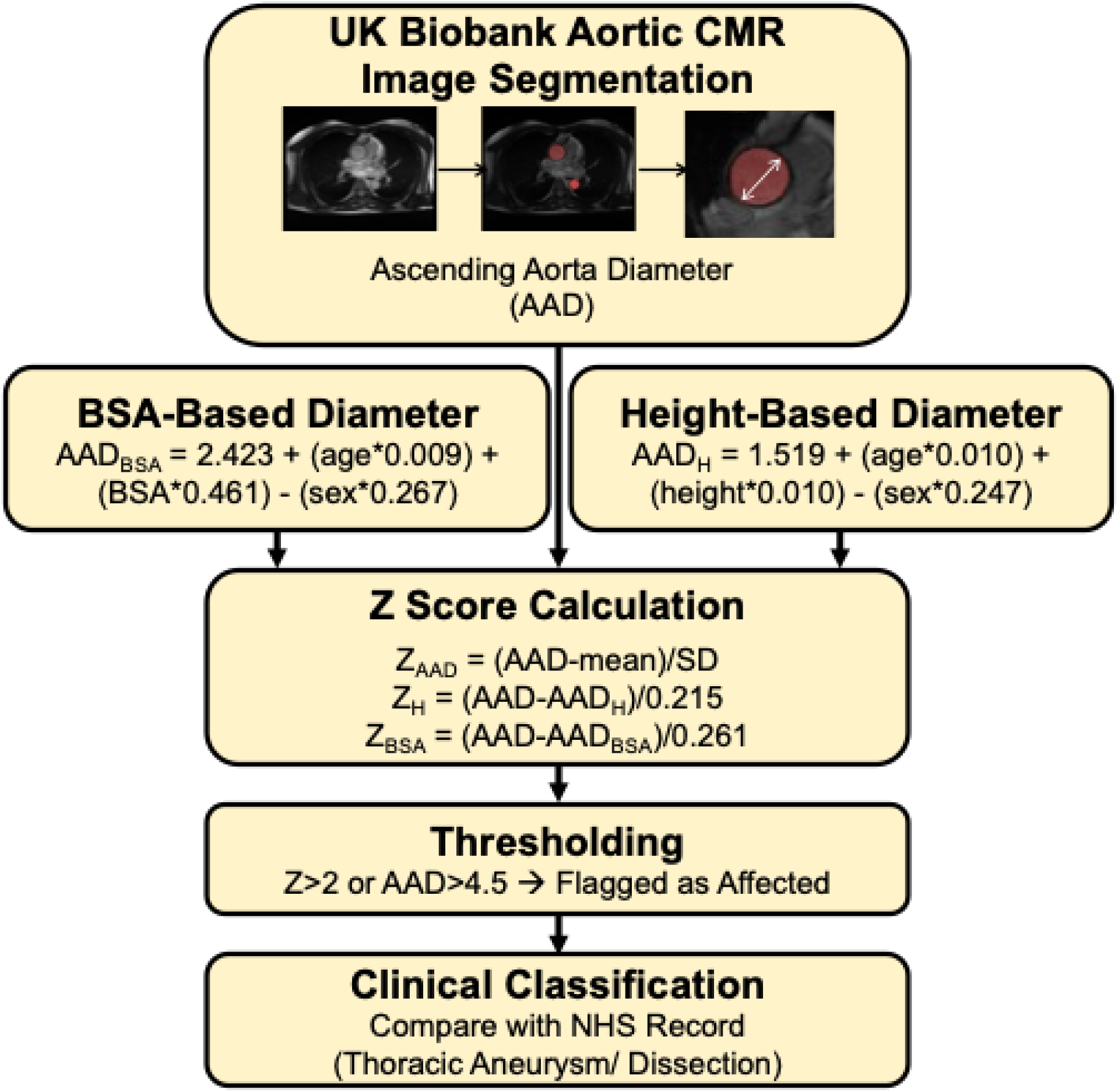
Flowchart of the study design, including AAD calculation and Z-score calculations for each approach.

### Z-Score Calculation and Thresholding

Three types of AAD Z-scores were calculated: (1) unadjusted AAD, (2) adjusted for BSA and (3) adjusted for height, in addition to age and sex.(8) The following equations from Devereux et. al. were used to calculate predicted AAD incorporating the effect of age and sex, and either BSA or height:

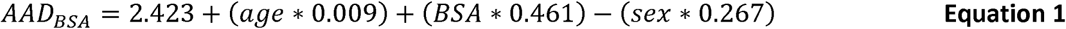

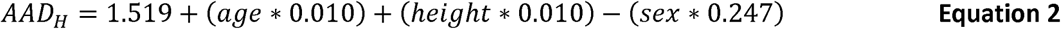

Z-scores were then calculated using following equations:

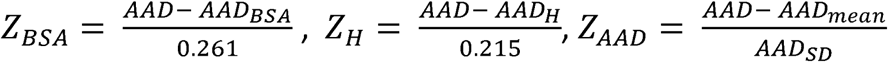

We compared the predictive performance of the single absolute AAD cut-off of 4.5 cm in the prediction of a later clinical diagnosis of thoracic aneurysm or dissection, with the three Z-scores: Z_AAD,_ Z_H_ and Z_BSA_. We classified individuals as affected if they had a linked National Health Service clinical record of either a thoracic aneurysm or a dissection, based on the list of clinical codes in Table 1 and used the R-based package ukbpheno to identify participants with a later clinical diagnosis of thoracic aneurysm or dissection.(16) We excluded participants who had the relevant clinical diagnosis before their imaging date.

**Table 1.** Thoracic Aorta Dissection/Dilatation/Surgery Identification Codes in the UK Biobank Cohort.

|  |  |
| --- | --- |
| **ICD10** |  |
| I71.1 | Thoracic aortic aneurysm, ruptured |
| I71.2 | Thoracic aortic aneurysm, without mention of rupture |
| **ICD9** |  |
| 4411 | Thoracic aneurysm, ruptured |
| 4412 | Thoracic aneurysm without mention of rupture |
| **OPCS3** |  |
| 321 | Repair of intra-thoracic aneurysm |
| 326 | Other plastic repair of intra-thoracic vessel |
| 329 | Other operations on intra-thoracic vessel |
| **OPCS4:** |  |
| L27.3 | Endovascular insertion of stent graft for thoracic aortic aneurysm |
| L28.3 | Endovascular insertion of stent for thoracic aortic aneurysm |

### Statistical Analysis

We evaluated the performance of different AAD cutoffs in predicting development of thoracic aortic dissection or aneurysm by calculating the detection rate (DR, the proportion of affected individuals who tested positive) at different pre-specified false positive rates (FPR). We compared performance of the corresponding test cutoffs with DR and FPR based on the guideline-endorsed absolute AAD cutoff of 4.5 cm. For comparison of groups, we used the Mann-Whitney U-test (for non-normally distributed variable) or Student’s t-test (for normally distributed variables) for continuous variables and the chi-squared test for categorical variables. Statistical significance was defined as p < 0.05. Continuous variables were reported as mean (standard deviation). All statistical analyses were conducted using Python (v3.9, https://www.python.org/) with the statsmodels and scikit-learn libraries.

## RESULTS

### Participant Demographics & Clinical Aneurysm/Dissection Cases

This study included 77,527 UKB participants, including 37,496 males and 40,031 females whose AAD was measured using aortic CMR imaging. Participant characteristics and differences by sex are provided in Table 2. Male participants were older, and taller, heavier, and had a higher BMI, BSA, DBP and SBP as compared to females, with values shown in Table 2. Average AAD was 3.3 (0.4) cm in the population. Aortic diameter in males was 3.4 (0.3) cm and 3.2 (0.3) cm in females (p<0.001).

**Table 2.** Demographic and clinical characteristics of the UK Biobank participants.

|  | **Overall**<br>**(N = 77,527)** | **Male**<br>**(N = 37,496)** | **Female**<br>**(N = 40,031)** | **p-value**<br>**(male vs**<br>**female)** |
| --- | --- | --- | --- | --- |
| Age (years) | 66.1 (8.0) | 66.7 (8.0) | 65.5 (7.9) | <0.001 |
| Height (cm) | 168.9 (9.2) | 175.7 (6.7) | 162.6 (6.3) | <0.001 |
| Weight (kg) | 75.8 (15.1) | 83.4 (13.4) | 68.6 (13.0) | <0.001 |
| BMI (kg/m<sup>2</sup>) | 26.5 (4.4) | 27.0 (3.9) | 26.0 (4.7) | <0.001 |
| BSA (m<sup>2</sup>) | 1.9 (0.2) | 2.0 (0.2) | 1.7 (0.2) | <0.001 |
| DBP (mmHg) | 80.5 (9.1) | 82.4 (8.7) | 78.7 (9.1) | <0.001 |
| SBP (mmHg) | 137.3 (16.8) | 140.6 (15.3) | 134.2 (17.5) | <0.001 |
| AAD (cm) | 3.3 (0.4) | 3.4 (0.3) | 3.2 (0.3) | <0.001 |

Seventy-two UKB imaging sub-study participants (0.1%; 57 males and 15 females) had a clinical thoracic aneurysm/dissection after their imaging date (median time from imaging to diagnosis was 852 days), and 77,455 participants were unaffected (Table 3). AAD was 4.2 (0.6) cm in the affected group and 3.3 (0.4) cm in the unaffected group. Height, weight and BSA were higher on average in the affected than unaffected group. The affected group also had higher age, SBP, DBP and BMI on average, but these differences were not statistically significant. Figure 2 plots the distribution of AAD affected individuals in red and unaffected individuals in blue. The majority of affected individuals (54, 44 males) had an aortic diameter below 4.5 cm and (69, 55 males) had an aortic diameter below 5.5 cm.

**Figure 2.**
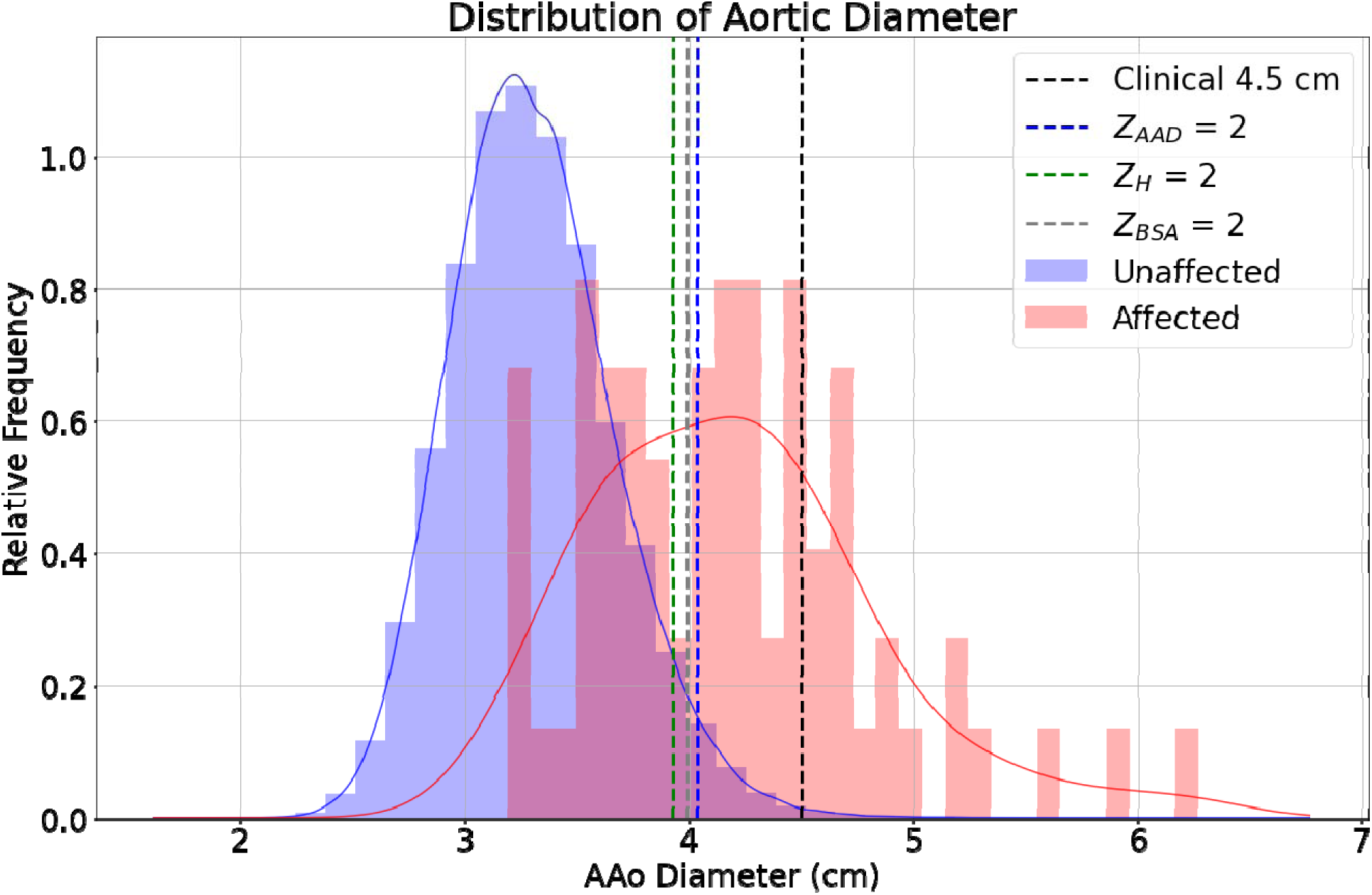
Histogram of representing AAD for participants in the whole cohort; red section represents clinically diagnosed aneurysm/dissection cases and blue represents the unaffected cases. Vertical lines represent the predictive threshold cutoff value of clinical 4.5 cm (black), Z_AAD_=2 (blue), Z_BSA_=2 (gray) and Z_H_=2 (green).

**Table 3.**
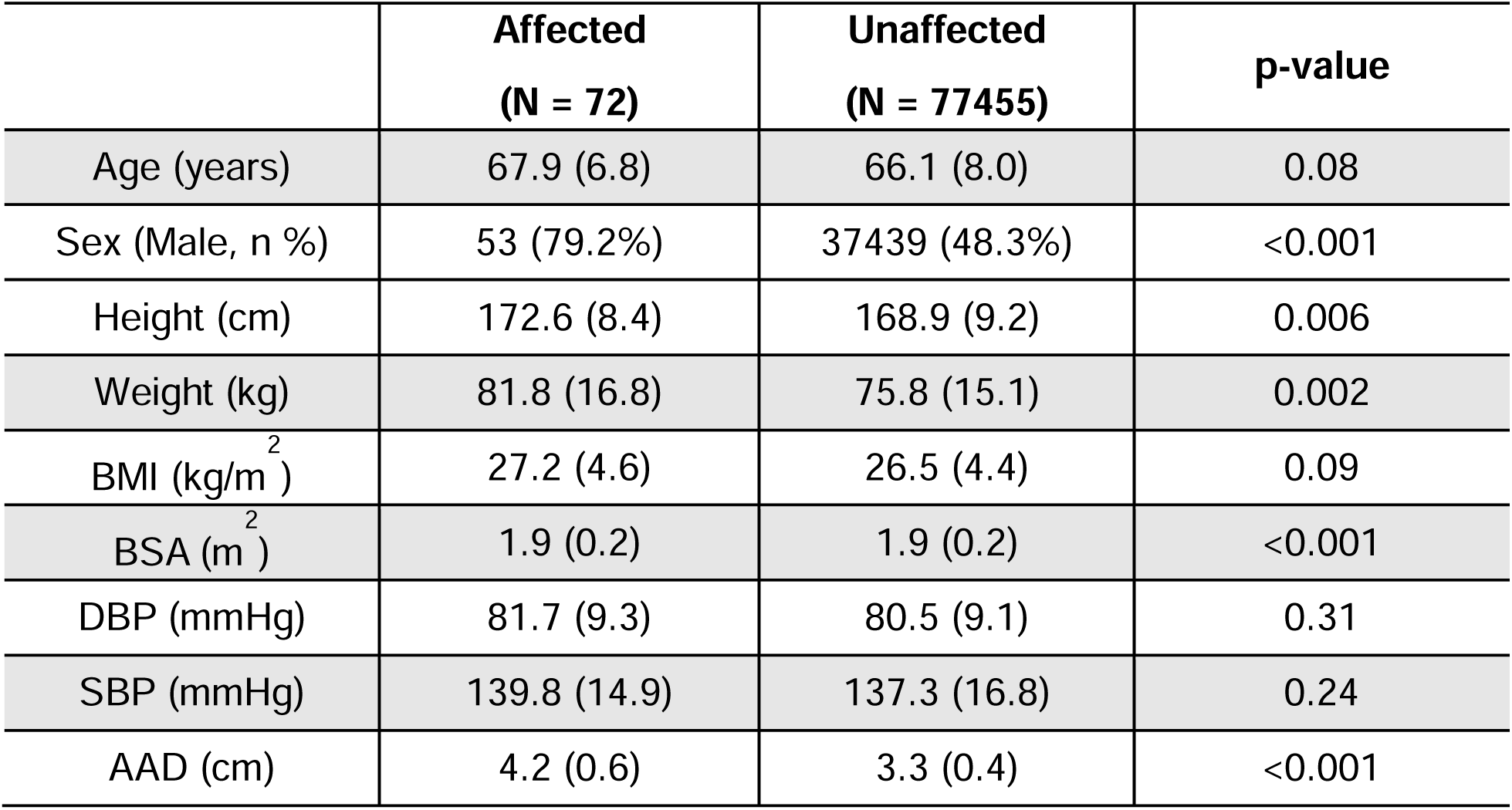
Comparison of demographic and clinical characteristics of individuals later affected or unaffected by thoracic aortic aneurysm or dissection.

|  | **Affected**<br>**(N = 72)** | **Unaffected**<br>**(N = 77455)** | **p-value** |
| --- | --- | --- | --- |
| Age (years) | 67.9 (6.8) | 66.1 (8.0) | 0.08 |
| Sex (Male, n %) | 53 (79.2%) | 37439 (48.3%) | <0.001 |
| Height (cm) | 172.6 (8.4) | 168.9 (9.2) | 0.006 |
| Weight (kg) | 81.8 (16.8) | 75.8 (15.1) | 0.002 |
| BMI (kg/m<sup>2</sup>) | 27.2 (4.6) | 26.5 (4.4) | 0.09 |
| BSA (m<sup>2</sup>) | 1.9 (0.2) | 1.9 (0.2) | <0.001 |
| DBP (mmHg) | 81.7 (9.3) | 80.5 (9.1) | 0.31 |
| SBP (mmHg) | 139.8 (14.9) | 137.3 (16.8) | 0.24 |
| AAD (cm) | 4.2 (0.6) | 3.3 (0.4) | <0.001 |

### Predictive performance of different AAD assessments

Of the 77,527 participants in the study, 224 (0.3%) had an AAD exceeding the guideline-endorsed cutoff of 4.5 cm; 18/72 affected and 224/77,455 unaffected individuals, giving a DR of 25% and FPR of 0.3%. Using an AAD Z-score of 2SD as a cutoff (corresponding to an aortic diameter of 4.0 cm), 2472/77,527 participants tested positive, 41/72 affected individuals and 2431/77,455 unaffected individuals, giving a DR of 57% and FPR of 3.1%. Using Z_BSA_ with a 2SD cutoff, 1922/77,527 tested positive, 34/72 affected individuals and 1888/77,455 unaffected individuals, yielding a DR of 47% and FPR of 2.4%. Using Z_H_ with 2SD cutoff, 2890/77,527 tested positive, 38/72 affected and 2852/77,455 unaffected individuals, yielding a DR of 53% and FPR of 3.7%.

Figure 3 shows the distribution of Z_AAD_, Z_H_ and Z_BSA_ values among affected and unaffected individuals and the corresponding DRs and FPRs at different Z-score values. Z_AAD_ (AUC=0.90, 95% CI = (0.86, 0.94)) provided better discrimination than Z_H_ (AUC=0.84, 95% CI = (0.78, 0.89)) and Z_BSA_ (AUC=0.83, 95% CI = (0.77, 0.88)).

**Figure 3.**
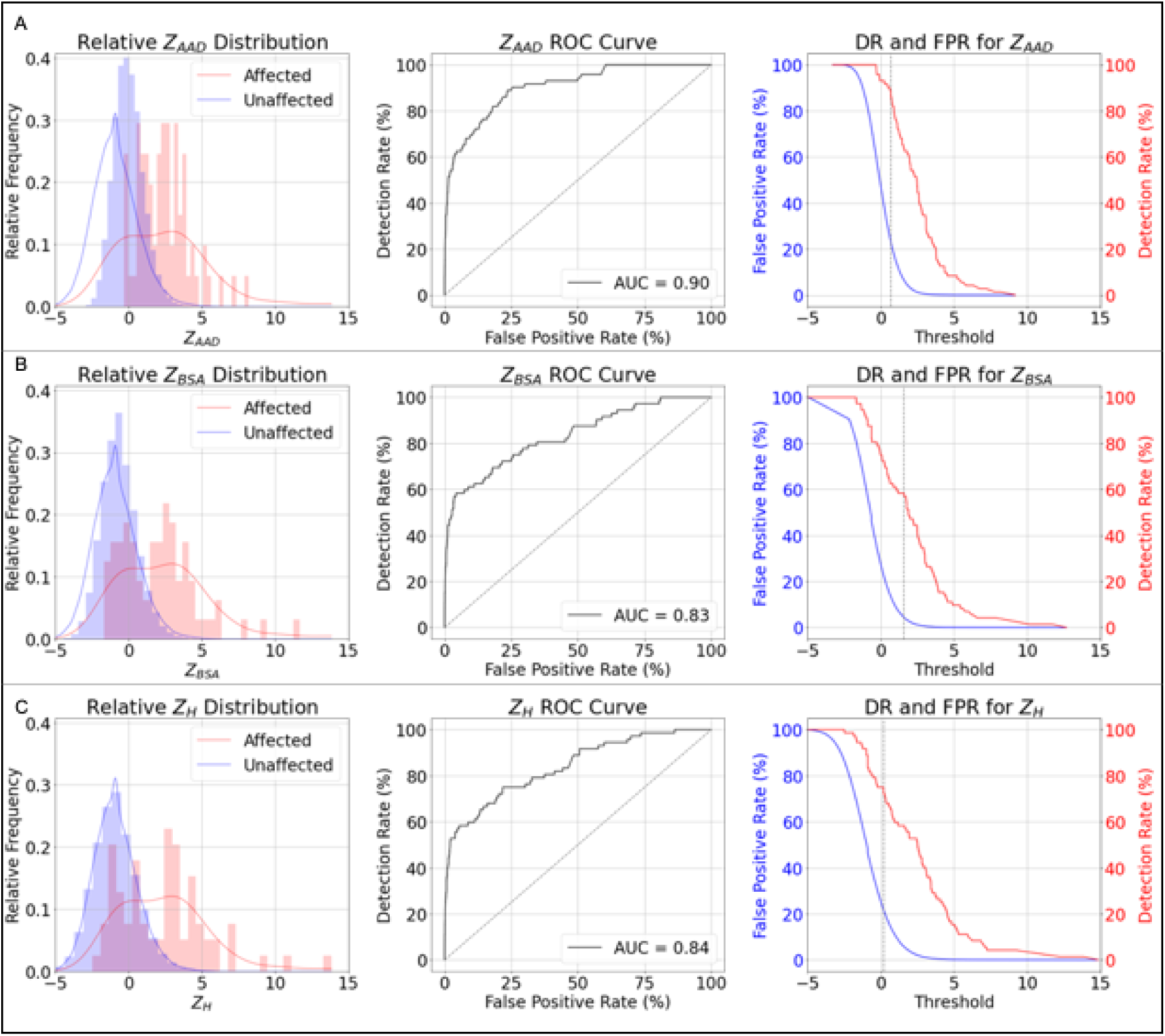
Performance of each predictive approach. Left panel plots the histogram of relative distribution of affected (red) and unaffected (blue) groups; middle panel plots the ROC curve with AUC inside the plot and the right panel plots DR (red) and FPR (blue) for each thresholding value plotted along the common Z-score axis for (A) Z_AAD_, (B) Z_BSA_ and (C) Z_H_, respectively.

Table 4 shows the DR at FPR cutoffs of 0.3%, 0.5%, 1%, 2%, 3%, 4% and 5% for Z_AAD_, Z_H_ and Z_BSA_. Z_AAD_ achieved the highest DR across all FPR cutoffs, with a DR of 25% at 0.3% FPR and 62.5% at 5% FPR. The values in parentheses denote the corresponding Z-score values (and in the case of Z_AAD_) the AAD measurement at each FPR cutoff.

**Table 4.** DR for thoracic aortic aneurysm or dissection at different FPRs (with corresponding Z-score cutoffs).

| **Predictor** | **Z<sub>AAD</sub>** | **Z<sub>BSA</sub>** | **Z<sub>H</sub>** |
| --- | --- | --- | --- |
| **0.3% FPR** | 25.0% (3.3) | 25.0% (3.3) | 25.0% (4.1) |
| **0.5% FPR** | 34.7% (3.0) | 26.4% (3.1) | 27.8% (3.8) |
| **1% FPR** | 40.3% (2.6) | 37.5% (2.6) | 36.1% (3.1) |
| **2% FPR** | 52.8% (2.3) | 45.8% (2.2) | 48.6% (2.5) |
| **3% FPR** | 55.6% (2.0) | 50.0% (1.9) | 52.8% (2.2) |
| **4% FPR** | 61.1% (1.9) | 56.9% (1.6) | 55.6% (1.9) |
| **5% FPR** | 62.5% (1.8) | 56.9% (1.6) | 56.9% (1.7) |

## DISCUSSION

We evaluated the predictive utility of current guideline-recommended absolute AAD thresholds for thoracic aorta management.(5–7) The 4.5 cm cutoff failed to identify three-quarters of individuals who subsequently developed thoracic aortic aneurysm or dissection, echoing prior registry data such as from IRAD.(2) In contrast, a simple population-normalized metric—Z_AAD_—substantially improved detection, identifying 57% of cases at Z>2, more than double the yield of the guideline-based method. This suggests that aortic size relative to age and sex-matched population norms is a more informative predictor of future risk than fixed thresholds or size adjustments based on body habitus.

Importantly, while Z_AAD_’s optimal cutoff yielded a high DR of 89%, it came with a 23% FPR—likely too high for widespread population screening. However, performance at lower FPRs was still clinically meaningful. At an FPR of 2%, which may be tolerable in targeted screening programs, Z_AAD_ detected 53% of cases—more than twice the detection achieved by the current 4.5 cm cutoff. The height-adjusted Z_H_ also showed balanced performance, identifying 56% of cases at a 4% FPR, suggesting it could serve as a confirmatory test following initial Z_AAD_-based screening. These results support a potential two-step strategy: initial screening using Z_AAD_ to maximize sensitivity, followed by more specific evaluation using adjusted metrics like Z_H_.

The observed sex-differences further underscore the need for individualized risk models. Despite near-equal sex distribution in the study population, most of aortic dissection and aneurysm events occurred in men.(17) We found that men had larger AADs than women even after accounting for BSA, supporting the development of sex-specific Z-score thresholds. Meanwhile, the affected group had higher BSA but similar BMI to the unaffected group, calling into question the utility of BMI-based adjustments in aortic risk prediction. These findings argue for prioritizing anatomical measurements and direct Z-score standardization over indirect anthropometric corrections.(18)

The integration of Z-score based aortic measurements with genetic risk factors offers a promising pathway toward personalised risk assessment. Polygenic risk scores and rare pathogenic variants in genes such as *FBN1*, *TGFBR1/2*, and *ACTA2* are increasingly recognised contributors to thoracic aortic disease.(19, 20) Combining imaging-derived Z-scores with genomic data could improve identification of individuals at high risk even before clinical manifestations, enabling tailored surveillance intervals and earlier surgical referral in selected patients. Future studies should test whether joint imaging–genetic models outperform current strategies in predicting incident aortic events, moving toward precision cardiovascular prevention.

There are also implications for clinical translation and scalability. Automated imaging analysis, including neural network–based segmentation, now enables extraction of aortic diameters and computation of Z-scores at scale.(15) This opens the possibility of embedding Z-score pipelines into routine cardiovascular MR or CT workflows, or even opportunistic case-finding when imaging is performed for other indications. Future work should also assess the cost-effectiveness of Z-score based surveillance, given the trade-off between improved sensitivity and potential increases in false positives, downstream imaging, and patient anxiety.

This study has a few limitations. First, our reliance on OPCS diagnostic codes likely reflects only diagnosed cases, and may miss individuals with asymptomatic or undiagnosed thoracic aortic dilation, particularly among women, who are historically less likely to receive an aortic diagnosis.(21) The apparent FPR may therefore be inflated, as individuals without a recorded diagnosis may nonetheless have undetected aortic dilation. Second, while our Z-scores were derived from high-resolution CMR, generalizability to echocardiography—more commonly used in clinical settings— remains to be established.(22) Future studies should aim to validate Z-score thresholds across high-risk populations and across ethnicities.

## CONCLUSION

Population-normalized Z-scores, particularly Z_AAD_, offer substantial improvements in the detection of individuals at risk of thoracic aortic aneurysm or dissection compared to current absolute diameter thresholds. Although higher false positive rates may limit their use in general screening, their sensitivity suggests utility in targeted clinical workflows, particularly for younger patients and women, who could be missed by existing guidelines. These findings support a shift toward individualized, data-driven approaches for management of thoracic aortic dissection and aneurysms.

## DATA AVAILABILITY STATEMENT

This research was conducted using data from the UK Biobank (Application Number 12113). The data used in this study are available to bona fide researchers upon application. Derived data generated in this study will be returned to the UK Biobank in accordance with their data sharing policy and will be made available to future approved researchers through the UK Biobank resource.

## REFERENCES

1. Meszaros I, Morocz J, Szlavi J, Schmidt J, Tornoci L, Nagy L, Szép L. Epidemiology and clinicopathology of aortic dissection. Chest. 2000;117(5):1271–8.

2. Pape LA, Tsai TT, Isselbacher EM, Oh JK, O’Gara PT, Evangelista A, et al. Aortic diameter≥ 5.5 cm is not a good predictor of type A aortic dissection: observations from the International Registry of Acute Aortic Dissection (IRAD). Circulation. 2007;116(10):1120–7.

3. Clouse WD, Hallett JW, Schaff HV, Spittell PC, Rowland CM, Ilstrup DM, Melton III LJ, editors. Acute aortic dissection: population-based incidence compared with degenerative aortic aneurysm rupture. Mayo Clinic Proceedings; 2004: Elsevier.

4. Chau KH, Elefteriades JA. Natural history of thoracic aortic aneurysms: size matters, plus moving beyond size. Progress in Cardiovascular Diseases. 2013;56(1):74–80.

5. Erbel R, Aboyans V, Boileau C, Bossone E, Di Bartolomeo R, Eggebrecht H, et al. 2014 ESC guidelines on the diagnosis and treatment of aortic diseases. European heart journal. 2014;35:2873–926.

6. Mazzolai L, Teixido-Tura G, Lanzi S, Boc V, Bossone E, Brodmann M, et al. 2024 ESC Guidelines for the management of peripheral arterial and aortic diseases: Developed by the task force on the management of peripheral arterial and aortic diseases of the European Society of Cardiology (ESC) Endorsed by the European Association for Cardio-Thoracic Surgery (EACTS), the European Reference Network on Rare Multisystemic Vascular Diseases (VASCERN), and the European Society of Vascular Medicine (ESVM). European Heart Journal. 2024:ehae179.

7. Members WC, Isselbacher EM, Preventza O, Hamilton Black III J, Augoustides JG, Beck AW, et al. 2022 ACC/AHA Guideline for the diagnosis and management of aortic disease: a report of the American Heart Association/American College of Cardiology Joint Committee on Clinical Practice Guidelines. Journal of the American College of Cardiology. 2022;80(24):e223–e393.

8. Devereux RB, de Simone G, Arnett DK, Best LG, Boerwinkle E, Howard BV, et al. Normal limits in relation to age, body size and gender of two-dimensional echocardiographic aortic root dimensions in persons≥ 15 years of age. The American journal of cardiology. 2012;110(8):1189–94.

9. van Kimmenade RR, Kempers M, de Boer MJ, Loeys BL, Timmermans J. A clinical appraisal of different Z-score equations for aortic root assessment in the diagnostic evaluation of Marfan syndrome. Genet Med. 2013;15(7):528–32.

10. Quezada E, Lapidus J, Shaughnessy R, Chen Z, Silberbach M. Aortic dimensions in Turner syndrome. American Journal of Medical Genetics Part A. 2015;167(11):2527–32.

11. Petersen SE, Matthews PM, Bamberg F, Bluemke DA, Francis JM, Friedrich MG, et al. Imaging in population science: cardiovascular magnetic resonance in 100,000 participants of UK Biobank-rationale, challenges and approaches. Journal of Cardiovascular Magnetic Resonance. 2013;15:1–10.

12. Sudlow C, Gallacher J, Allen N, Beral V, Burton P, Danesh J, et al. UK biobank: an open access resource for identifying the causes of a wide range of complex diseases of middle and old age. PLoS Medicine. 2015;12(3):e1001779.

13. DuBois D. A formula to estimate the approx-imate surface area if height and weight be known. Arch intern med. 1916;17:863–71.

14. Bai W, Suzuki H, Qin C, Tarroni G, Oktay O, Matthews PM, Rueckert D, editors. Recurrent neural networks for aortic image sequence segmentation with sparse annotations. Medical Image Computing and Computer Assisted Intervention–MICCAI 2018: 21st International Conference, Granada, Spain, September 16-20, 2018, Proceedings, Part IV 11; 2018: Springer.

15. Bai W, Sinclair M, Tarroni G, Oktay O, Rajchl M, Vaillant G, et al. Automated cardiovascular magnetic resonance image analysis with fully convolutional networks. Journal of Cardiovascular Magnetic Resonance. 2018;20(1):65.

16. Yeung MW, van der Harst P, Verweij N. ukbpheno v1. 0: An R package for phenotyping health-related outcomes in the UK Biobank. STAR protocols. 2022;3(3):101471.

17. Olsson C, Thelin S, Sta hle E, Ekbom A, Granath F. Thoracic aortic aneurysm and dissection: increasing prevalence and improved outcomes reported in a nationwide population-based study of more than 14 000 cases from 1987 to 2002. Circulation. 2006;114(24):2611–8.

18. de Simone G, Daniels SR, Devereux RB, Meyer RA, Roman MJ, de Divitiis O, Alderman MH. Left ventricular mass and body size in normotensive children and adults: assessment of allometric relations and impact of overweight. Journal of the American College of Cardiology. 1992;20(5):1251–60.

19. Aragam KG, Jiang T, Goel A, Kanoni S, Wolford BN, Atri DS, et al. Discovery and systematic characterization of risk variants and genes for coronary artery disease in over a million participants. Nature genetics. 2022;54(12):1803–15.

20. Dietz HC, Cutting CR, Pyeritz RE, Maslen CL, Sakai LY, Corson GM, et al. Marfan syndrome caused by a recurrent de novo missense mutation in the fibrillin gene. Nature. 1991;352(6333):337-9.

21. Chung J, Coutinho T, Chu MW, Ouzounian M. Sex differences in thoracic aortic disease: a review of the literature and a call to action. The Journal of Thoracic and Cardiovascular Surgery. 2020;160(3):656–60.

22. Saura D, Dulgheru R, Caballero L, Bernard A, Kou S, Gonjilashvili N, et al. Two-dimensional transthoracic echocardiographic normal reference ranges for proximal aorta dimensions: results from the EACVI NORRE study. European Heart Journal-Cardiovascular Imaging. 2017;18(2):167–79.

